# The genetic overlap between Alzheimer’s disease, amyotrophic lateral sclerosis, Lewy body dementia, and Parkinson’s disease

**DOI:** 10.1101/2022.09.26.22280159

**Authors:** Douglas P. Wightman, Jeanne E. Savage, Elleke Tissink, Cato Romero, Iris E. Jansen, Danielle Posthuma

## Abstract

Neurodegenerative diseases are a group of disorders characterised by neuronal cell death causing a variety of physical and mental problems. While these disorders can be characterised by their phenotypic presentation within the nervous system, their aetiologies differ to varying degrees. Some disorders, such as Lewy body dementia and Parkinson’s disease, show overlap in the major proteins found in aggregates, and some diseases, like Alzheimer’s disease, amyotrophic lateral sclerosis, and Parkinson’s disease, are influenced by the same non-neuronal cell types (microglia), suggesting partly shared aetiologies. The identification of shared genetic risk factors common to many neurodegenerative diseases may highlight fundamental biological processes involved in neurodegeneration and provide promising targets for treatment and drug repurposing. The majority of genetic evidence for overlap between neurodegenerative diseases has been pairwise, with little genetic evidence for genes or biological processes found across more than two neurodegenerative diseases. In this study, we aimed to identify overlap between the four investigated neurodegenerative disorders (Alzheimer’s disease, amyotrophic lateral sclerosis, Lewy body dementia, and Parkinson’s disease) at the variant, gene, genomic locus, gene-set, cell, or tissue level, with specific interest in overlap between three or more diseases. Using local genetic correlation, we found that the *TMEM175* locus was a shared locus between amyotrophic lateral sclerosis, Lewy body dementia, and Parkinson’s disease, and the HLA region was shared between Alzheimer’s disease, amyotrophic lateral sclerosis, and Parkinson’s disease. We also highlighted genes, genomic loci, gene-sets, cell types, and tissue types which may be important to two or more disorders by analysing the association of variants with a common factor estimated from the four disorders. Our study successfully highlighted genetic loci and tissues associated with two or more neurodegenerative diseases.

## Introduction

Neurodegenerative diseases are a group of late-onset disorders characterised by neuronal cell death leading to movement, cognitive and/or behavioural problems^1^. Most neurodegenerative diseases can be characterised into four groups defined by the typical protein identified in the aggregates of affected patients; (1) Amyloidoses, like Alzheimer’s disease (AD), are defined by the presence of amyloid proteins, (2) Tauopathies, like chronic traumatic encephalopathy, are defined by tau proteins, (3) Synucleinopathies, like dementia with Lewy bodies (DLB), are defined by α-synuclein, and (4) TAR DNA-binding protein 43 (TDP-43) proteinopathies, like amyotrophic lateral sclerosis (ALS), are defined by TDP-43. Despite this categorisation, more than one of these proteins can be a risk factor in a single neurodegenerative disease. Parkinson’s disease (PD) is classified as a synucleinopathy due to accumulations of α-synuclein, but tau is increasingly being identified as genetic risk factor in PD^2^. Similarly, AD is identified as an amyloidosis but tau is also present in the aggregates^1^. The same proteins can be risk factors for multiple neurodegenerative diseases which implies overlap at a subcellular level between different neurodegenerative diseases. The structure of protein aggregates and the cells involved with the disease process can also overlap between diseases. For example, both PD and DLB present with Lewy body aggregates^2,3^ and microglia are implicated in both PD, ALS and AD^1,2,4^ These findings suggest some degree of overlap in some neurodegenerative diseases outside of neuronal loss.

Previous studies have looked into the genetic overlap between varying numbers of neurodegenerative diseases (**Table 1**). Arneson *et al*. (2018)^5^ searched the GWAS catalog^6^ (19 September 2017) for variants and genes that overlap between AD, ALS, and PD and found that no significant variants or genes overlapped between the three traits. They performed pathway enrichment analysis for the identified genes and found 1 gene-set, related to vesicle mediated transport, which was significant in all 3 traits. Despite only finding a single overlapping pathway between all three diseases, they did find 2 genes and 10 pathways that overlapped between AD and PD. This suggests that AD and PD shared more genetic risk than ALS with either of AD or PD, but this may be partly due to the lower sample size of ALS GWAS available in September 2017 compared to AD or PD. Using GWAS summary statistics, Karch *et al*. (2018)^7^ also looked at ALS and a series of other neurodegenerative diseases and failed to find overlap of ALS^8^ risk variants (12,577 cases and 23,475 controls) with PD^9^ (5,333 cases and 12,019 controls) and AD^10^ (17,008 cases and 37,154 controls). Van Rheenen *et al*. (2021)^11^ (ALS: 27,205 cases and 110,881 controls) found significant genetic correlations between ALS and AD^12^ (rg=0.31, SE=0.12, P=9.6×10^−3^) and ALS and PD^13^ (rg=0.16, SE=0.061, P=0.011). They also found evidence for shared loci (HLA and *GAK/TMEM175*) between ALS and PD and a shared locus between AD and ALS (*TSPOAP1-AS1*).

**Table 1:** The genetic overlap between Alzheimer’s disease (AD), amyotrophic lateral sclerosis (ALS), Lewy body dementia (LBD) (or dementia with Lewy bodies (DLB)), and Parkinson’s disease (PD) in previous studies^5,11,14–16^.

| Trait pairs | Summary of previous findings |
| --- | --- |
| AD-ALS | Arneson *et al.* (2018) identified one shared pathway (vesicle mediated transport); Van Rheenen *et al.* (2021) found global genetic correlation of 0.31 (0.12) and a one shared locus (*TSPOAP1-AS1*). |
| AD-LBD/DLB | Chia *et al.* (2021) identified the *APOE* and *BIN1* loci as shared; Guerreiro *et al.* (2016) found a global genetic correlation of 0.578 (0.075). |
| AD-PD | Arneson *et al.* (2018) identified 10 shared pathways, and 2 shared genes (*HLA-DRB1* and *MAPT*); Guerreiro *et al.* (2016) found a global genetic correlation of 0.08 (0.101); Stolp Andersen *et al.* (2022) identified a significant local genetic correlation at the HLA locus. |
| ALS-LBD/DLB | NA |
| ALS-PD | Arneson *et al.* (2018) identified one shared pathway (vesicle mediated transport); Van Rheenen *et al.* (2021) found global genetic correlation of |
|  | 0.16 (0.061) and two shared loci (HLA and *GAK/TMEM175*). |
| LBD-PD | Chia *et al.* (2021) identified the *GBA*, *TMEM175*, and *SNCA* loci as shared; Guerreiro *et al.* (2016) found a global genetic correlation of 0.362 (0.107). |

Chia *et al*. (2021)^14^ performed a GWAS of Lewy body dementia (LBD) (2591 cases and 4027 controls), a wider category of dementia which includes DLB and PD with dementia, and found that polygenic risk models trained from AD^12^ and PD^13^ GWAS summary statistics were significantly associated with LBD status, suggesting shared genetic mechanisms. They also found loci associated with LBD at known AD (*APOE* and *BIN1*) and PD (*GBA, TMEM175* and *SNCA*) loci. Guerreiro *et al*. (2016)^15^ found a significant genetic correlation between AD (959 cases and 1403 controls) and DLB (788 cases and 1403 controls), and PD (804 cases and 1403 controls) and DLB, but not between AD and PD (AD-DLB: rg=0.578, SE=0.075, P=1.1×10^−12^; PD-DLB: rg=0.362, SE=0.107, P=7.1×10^−4^; AD-PD, rg=0.08, SE=0.101, P=0.39). However, Stolp Andersen *et al*. (2022)^16^ found a significant local genetic correlation between AD^17^ (24,087 cases, 47,793 proxy-cases, and 383,378 controls) and PD^13^ (37,688 cases, 18,618 proxy-cases, and 1,417,791 controls) at the HLA region. These previous studies suggest a degree of overlap between AD, ALS, LBD, and PD at the global or local level, with the exception of LBD and ALS where we were unable to find a study explicitly assessing overlap between these two traits.

In this study, we aim to quantify the degree of genetic overlap between 4 chosen neurodegenerative traits (AD, ALS, LBD, and PD). We chose to study AD, ALS, LBD, and PD because they have varying degrees of global genetic correlation, are characterised by different protein aggregates, and have been included in relatively large sample size GWAS. We assessed the degree of genetic overlap at the variant, gene, locus, gene-set, cell, and tissue level. We utilised summary statistics from Wightman *et al*. (2021)^18^ (AD: 39,918 cases and 358,140 controls), van Rheenen *et al*. (2021)^11^ (ALS: 27,205 cases and 110,881 controls), Chia *et al*. (2021)^14^ (LBD: 2,591 cases and 4,027 controls), and Nalls *et al*. (2019)^13^ (PD: 15,056 cases, 12,637 controls, 18,618 proxy cases, and 436,419 proxy controls). All of the summary statistics were generated from individuals of European ancestry. We estimated a genomicSEM^19^ common factor model to identify the association of variants with a common factor derived from all four neurodegenerative traits. We then used the resulting variant associations with the common factor to investigate genes, loci, gene-sets, cell types, and tissue types enriched in association signal. We aimed to identify overlap between all four traits to highlight biological process and genetic risk factors important to neurodegeneration because common biological processes and genetic risk factors may be useful targets for drug development and repurposing. We also looked at the variant associations within each trait individually and compared the findings across the input traits to determine if the common factor model was able to help identify variants, loci, gene-sets, cell types, and tissues types that were shared between one or more traits that were not observable by comparing the individual trait level results. We then investigated the local genetic correlation between the four traits at loci identified from the common factor using LAVA^20^.

## Results

### LDSC Analyses

We first estimated the heritability on the liability scale for all four traits using LDSC regression^21^. The heritability estimates of the input datasets were relatively low: AD: h2_liability_=0.053 (SE=0.010); ALS: h2_liability_=0.016 (SE=0.0017); LBD: h2_liability_=0.10 (SE=0.044); PD: h2_liability_=0.034 (SE=0.0031) (**Supplementary Table 1**). These estimates were similar to the heritability estimates reported in the original studies (**Supplementary Table 1**), except PD where the original paper did not report the heritability estimate when including the proxy dataset. The heritability estimate reported in GWAS Atlas^22^ from the data derived from a preprint version of the original PD study, which did include the proxy dataset, was similar to the estimate we identified (h2_liability_=0.022). To explore the global genetic correlation between AD, ALS, LBD, and PD we performed pairwise LDSC regression genetic correlation analysis (**Figure 1**).

**Figure 1:**
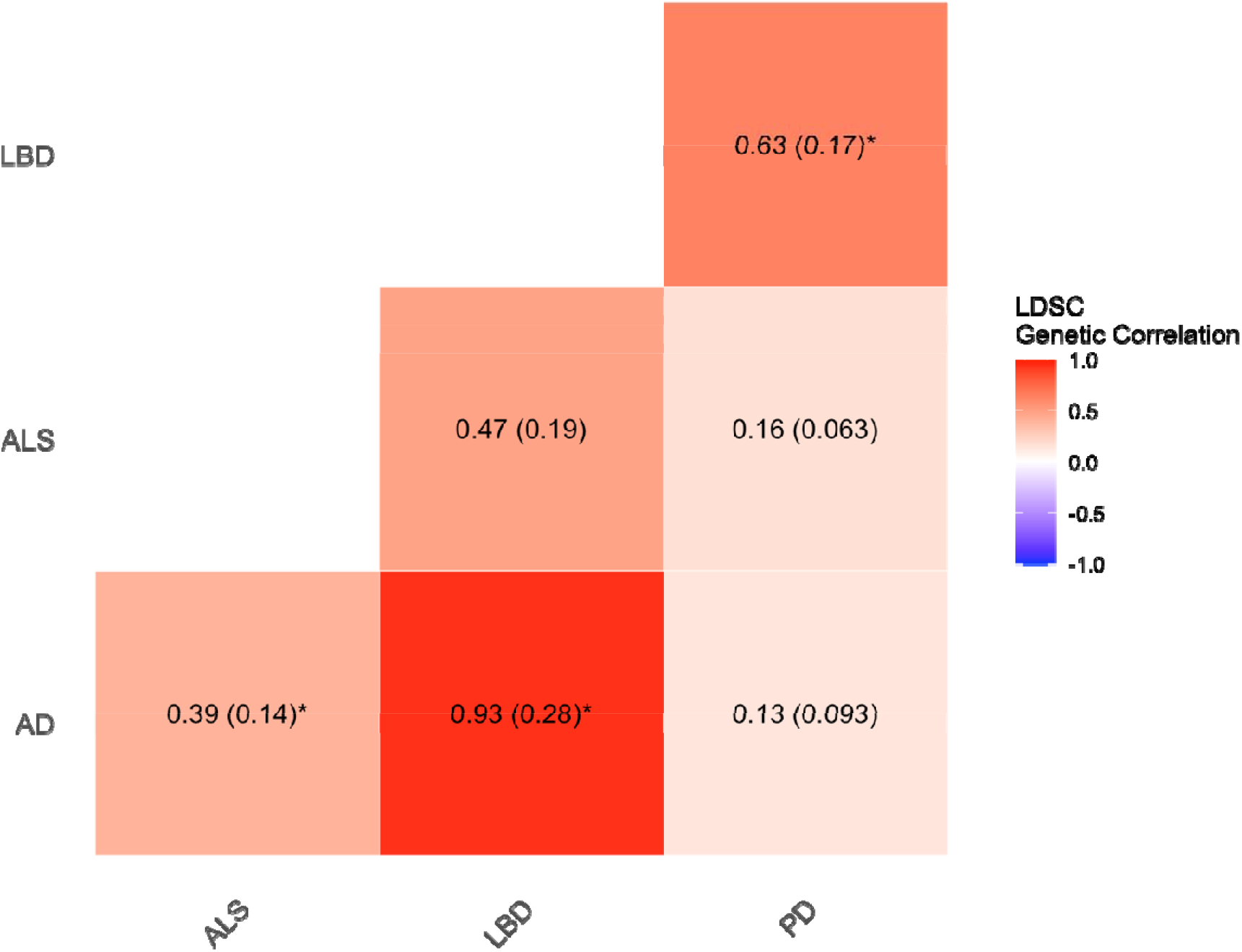
LDSC regression genetic correlations between AD, ALS, LBD, and PD identifies a strong genetic correlation between AD and LBD, moderate correlations between AD and ALS, ALS and LBD, and PD and LBD, weak correlations between ALS and PD, and AD and PD. Significant correlations after Bonferroni correction for 6 genetic correlation tests are highlighted by an asterisk. The colour of each block represents the genetic correlation value (rg) and standard error estimates are included in brackets.

The results indicated a high genetic correlation between AD and LBD (rg=0.93, SE=0.28, P=8.0×10^−4^), a moderate genetic correlation between AD and ALS (rg=0.39, SE=0.14, P=0.0036), LBD and ALS (rg=0.47, SE=0.19, P=0.011), and LBD and PD (rg=0.62, SE=0.17, P=2.0×10^−4^), a low genetic correlation between ALS and PD (rg=0.16, SE=0.063, P=0.011) and no nominally significant genetic correlation between AD and PD (rg=0.13, SE=0.092, P=0.15). All correlations, apart from between AD and PD, were nominally significant, but after Bonferroni correction for 6 tests, only the genetic correlations between AD and LBD, AD and ALS, and LBD and PD were significantly different from 0. The genetic correlation between AD and LBD was robust to the exclusion of the larger *APOE* region (GRCh37: 19-40000000-50000000) (rg=0.87, SE=0.21, P=2.34×10^−5^), all other genetic correlations were also largely unaffected by the exclusion of the APOE region (**Supplementary Table 1**).

### GenomicSEM Common Factor Model

#### Common Factor Model Results

To look for genetic variants important to AD, ALS, LBD, and PD, we fit a common factor model to all four traits using genomicSEM. Due to the low effective sample size of the LBD data (N_eff_=6,306.41), we specified in the model that LBD should load fully on the common factor (0 residual variance) to avoid a Heywood case where LBD would have negative residual variance. The model was also specified so that the variance of the common factor was 1. The resulting model fit the data relatively well (Chisq=8.94, df=3, P_chisq=0.03, CFI=0.92, SRMR=0.16) (**Supplementary Table 2**). The factor loadings (**Figure 2**) of each of the traits show that LBD, as specified, loads entirely onto the common factor (standardised estimate=1.20, standardised SE=0.16), with AD showing the second highest factor loading (standardised estimate=0.61, standardised SE=0.10), followed by ALS (standardised estimate=0.48, standardised SE=0.10), and PD (standardised estimate=0.35, standardised SE=0.07). We also tested a model where the LBD residual variance could have any value larger 0 and this did not change the model for the other traits and only slightly adjusted the LBD parameters. This model had a slightly worse fit (Chisq=8.94, df=3, P_chisq=0.011, CFI=0.91, SRMR=0.16) so we chose to continue with the model where LBD was specified to have a residual variance of 0.

**Figure 2:**
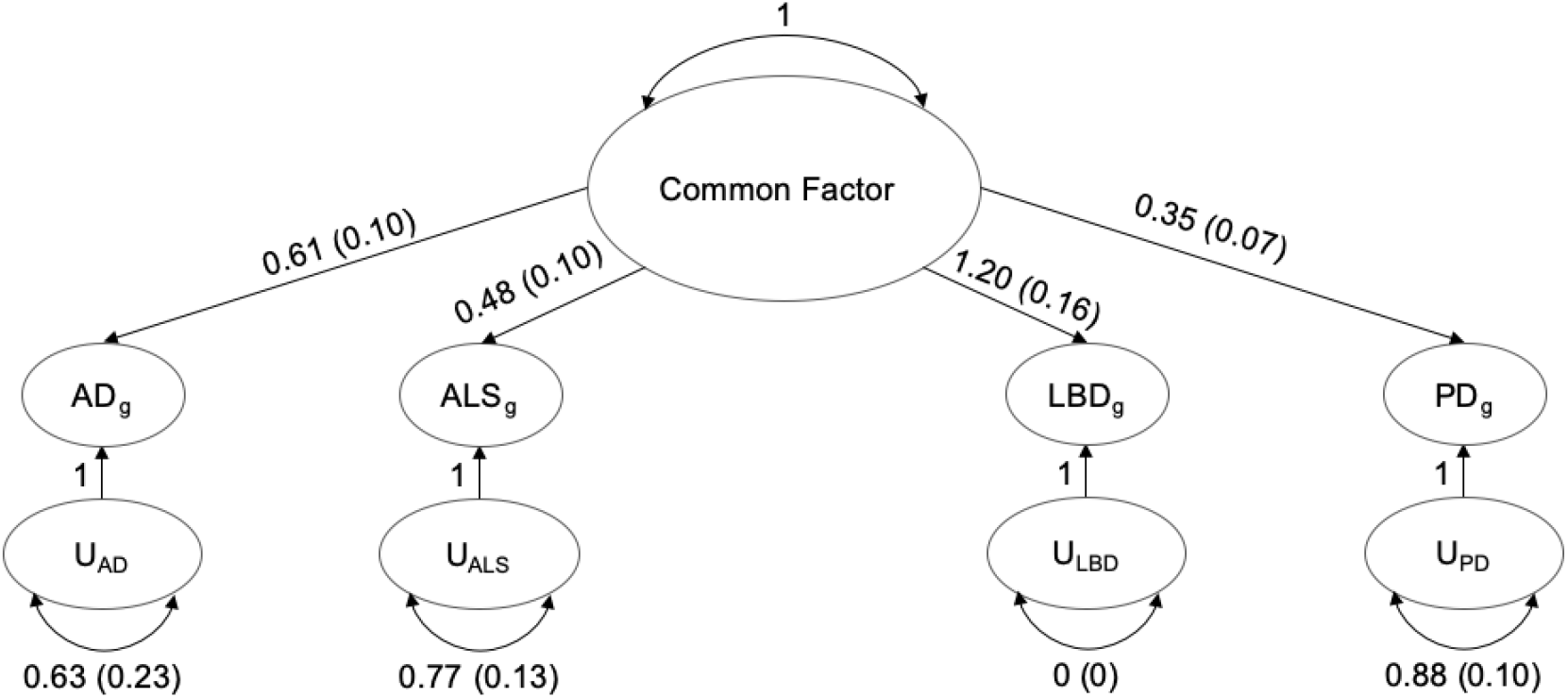
The factor loadings of the 4 neurodegenerative traits on the common factor indicate the highest loading of LBD, then AD, ALS and PD. Residual variance is represented by circular arrows. Factor loadings are represented by straight arrows. Values are indicated next to the arrows and standard errors are contained in brackets. All factor loadings are significantly different from 0.

We then used this common factor model to estimate the associations between the variants with the common factor. Only variants present in all 4 of the input GWAS were included. This resulted in association estimates for 5,951,489 genetic variants (**Figure 3**). The model was unable to obtain results from 690 variants due to computational limitations. The 690 failed variants were all in regions which were significant in at least one of the input traits (AD: *CR1, MS4A6A, ABCA7, APOE*; ALS: *C9orf72, G2E3*; LBD: *APOE*; PD: *SNCA, NDUFAF2*). In all of these regions, there were other variants which were significantly or suggestively associated with the common factor, so these regions were still highlighted even with the loss of these variants.

**Figure 3:**
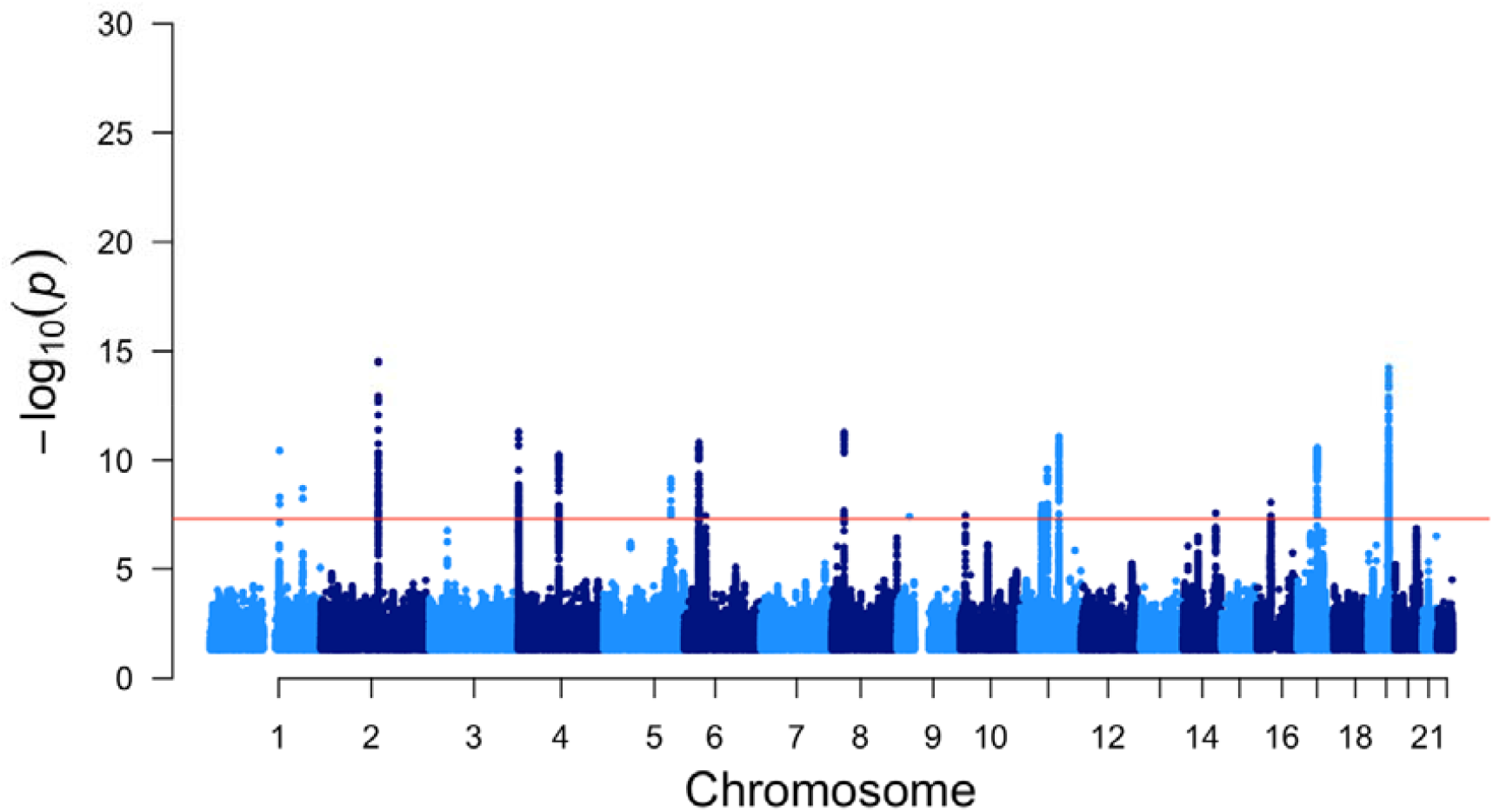
Manhattan plot of the variants included in the common factor model analysis identifies 2676 significant variants (P<5×10^−8^).

We identified 2,676 variants significantly associated with the common factor (P<5×10^−8^). The common factor association results had a genomic inflation factor of 1.14. When restricted to the same variants as the common factor, the association results in all 4 input traits had lower genomic inflation factors than the common factor association results (AD: 1.09; ALS: 1.10; LBD: 1.004; PD: 1.08). The LDSC intercept for the common factor (assuming total sample size of 362,647.1) was 1.0041 (SE=0.013) and the ratio was 0.026 (0.084), which suggests the majority of the inflation was due to polygenicity. There were more variants significantly associated with the common factor than with 3 of the input traits (AD: 1,591; ALS: 160; LBD: 83; PD: 2,733). There were 25 variants that were significantly associated with the common factor but not with any of the input traits; however, all of these variants were distributed across regions which already contained other significant variants in at least one input trait (*TMEM175*, HLA, *TNIP1*, and *MAPT* regions).

#### Common Factor Loci

We used FUMA (v1.3.8) to identify significant genomic loci from the common factor summary statistics and the summary statistics of the 4 input traits to identify genomic regions associated with AD, ALS, LBD, and PD. After merging loci less than 100Kb apart, there were 57 unique significant loci identified by FUMA across the common factor and the 4 input traits (**Supplementary Table 3**). Of those 57 loci, 18 were significant in the common factor, 25 were significant in AD, 10 were significant in ALS, 7 were significant in LBD, and 23 were significant in PD. All of the loci identified from the common factor were significant in 1 or more input traits and all loci that were significant in two or more input traits were significant in the common factor. The common factor model did not identify any additional loci. This suggests that the common factor did not miss any loci that would have been identified from the separate 4 input traits but did not identify any additional loci not discoverable from the input traits. Seven loci were identified as significant in two or more of the input traits and all of these loci were significant in the common factor (**Supplementary Table 4**). The HLA locus was significantly associated with AD, ALS, PD, and the common factor. The *BIN1* and *APOE* loci were significantly associated with AD, LBD, and the common factor. The *GBA, SNCA*, and *TMEM175* loci were significantly associated with LBD, PD, and the common factor. The *TNIP1* locus was significantly associated with AD, ALS, and the common factor (**Figure 4**). With the exception of the HLA locus, all the relationships between the input traits were pairwise.

**Figure 4:**
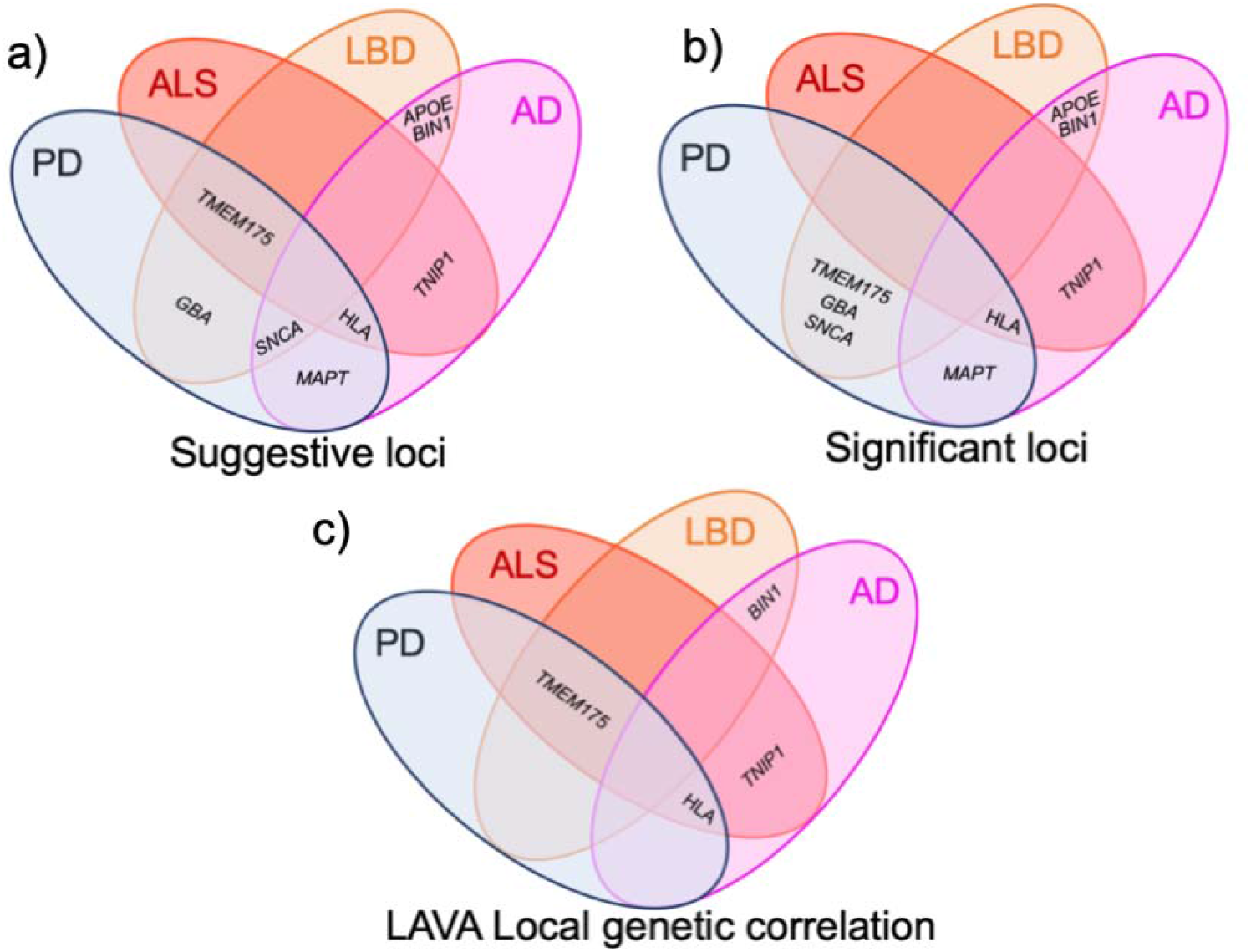
Venn diagrams showing the overlap between AD, ALS, LBD, and PD. A) The overlap of significant loci (P<5×10^−8^), defined by FUMA, across the 4 input traits. B) The overlap of suggestive loci (P<1×10^−5^), defined by FUMA, across the 4 input traits. C) The regions with significant local genetic correlation after Bonferroni correction for the number of genetic correlations performed in LAVA.

To determine the contribution of the input traits to the association of the lead variants identified from the common factor, we looked at the P-values of the common factor lead variants in the summary statistics of the input traits. The association signal distribution of the lead variants identified from the common factor reflects the factor loadings from the common factor model, with P-values of the common factor lead variants being lower in AD and LBD compared to ALS and PD (**Supplementary Figure 1**; **Supplementary Table 5**). We also explored whether there were three-way or four-way relationships at loci just below the significance threshold by repeating the FUMA loci definition as described above, except we allowed for loci to be defined around variants with suggestive P-values (P<1×10^−5^) (**Supplementary Table 3**; **Supplementary Note**). From this analysis, we still did not identify any loci common to all four input traits but did find that *TMEM175* was suggestive in ALS as well as significant in LBD, PD, and the common factor, and that *SNCA* was suggestive in AD as well as significant in LBD, PD, and the common factor (**Supplementary Note**; **Figure 4**).

#### Common Factor Gene Analysis

We performed MAGMA gene analysis implemented in FUMA to identify genes associated with the common factor and the 4 input traits. After Bonferroni correction for the number of genes tested within each trait, we identified 77 genes associated with the common factor, 73 associated with AD, 26 associated with ALS, 6 associated with LBD, and 61 associated with PD (**Supplementary Table 6**). Seven of the genes significantly associated with the common factor were not significantly associated with any of the input traits. Four of these genes were present in loci identified as significant in one or more of the input traits; one gene (*FGFRL1*) was in the *TMEM175* locus, two genes (*HLA-DQB1* and *C6orf10*) were in the HLA region, 1 gene (*PRR14*) was on the edge of a locus identified as a significant locus in PD. Three genes (*DOC2A, PPP4C*, and *ALDOA*) were in a locus not identified as a significant locus by FUMA in any of the input traits or the common factor. However, the *DOC2A* region was significantly associated with AD in a previous GWAS^23^. In our study, there was some association signal with this gene (*DOC2A*) in AD and LBD (P_CF_=1.65×10^−6^, P_AD_=2.17×10^−4^, P_ALS_=0.21, P_LBD_=2.52×10^−4^, P_PD_=0.10) suggesting that this gene may be present in a general dementia risk locus. There were similar association signals in the other two genes in this locus (**Supplementary Table 6**).

#### Common Factor Gene-set Analysis

In order to investigate how the genetic variants associated with the common factor aggregate together at different levels, we used FUMA and MAGMA to perform gene-set analyses using gene-sets defined by tissue and cell gene expression, and genes known to contribute to biological processes. On the tissue level, 10 GTEx tissue gene-sets were significantly associated with the common factor association signal after Bonferroni correction for 54 tested tissues (**Figure 5**; **Supplementary Table 7**). All 10 of these tissues were brain tissues and only 2 of these tissues were more significantly associated with an input dataset than the common factor (Brain Cerebellum in ALS, and Brain Cerebellar hemisphere in ALS and PD). The associations appear to be largely driven by ALS and PD genetics, with AD and LBD not even being nominally significant in any of these 10 tissues. Brain Putamen basal ganglia, brain hippocampus, brain anterior cingulate cortex BA24, and brain amygdala were significant in the common factor but not significant in any of the input traits. After conditioning on the top associated tissue (brain nucleus accumbens basal ganglia), none of the significantly associated tissues remained significant. This suggests that the common factor association signal was distributed in genes which are relevant to brain tissue in general rather than any specific tissue. All tissues that were significant in one of the input traits were significant in the common factor, except whole blood and spleen which were only significant in AD. These results suggest that genes differentially expressed in brain tissue were associated with the common factor largely due to the contribution of ALS and PD, and the association was with brain tissue as a whole rather than specific brain tissues.

**Figure 5:**
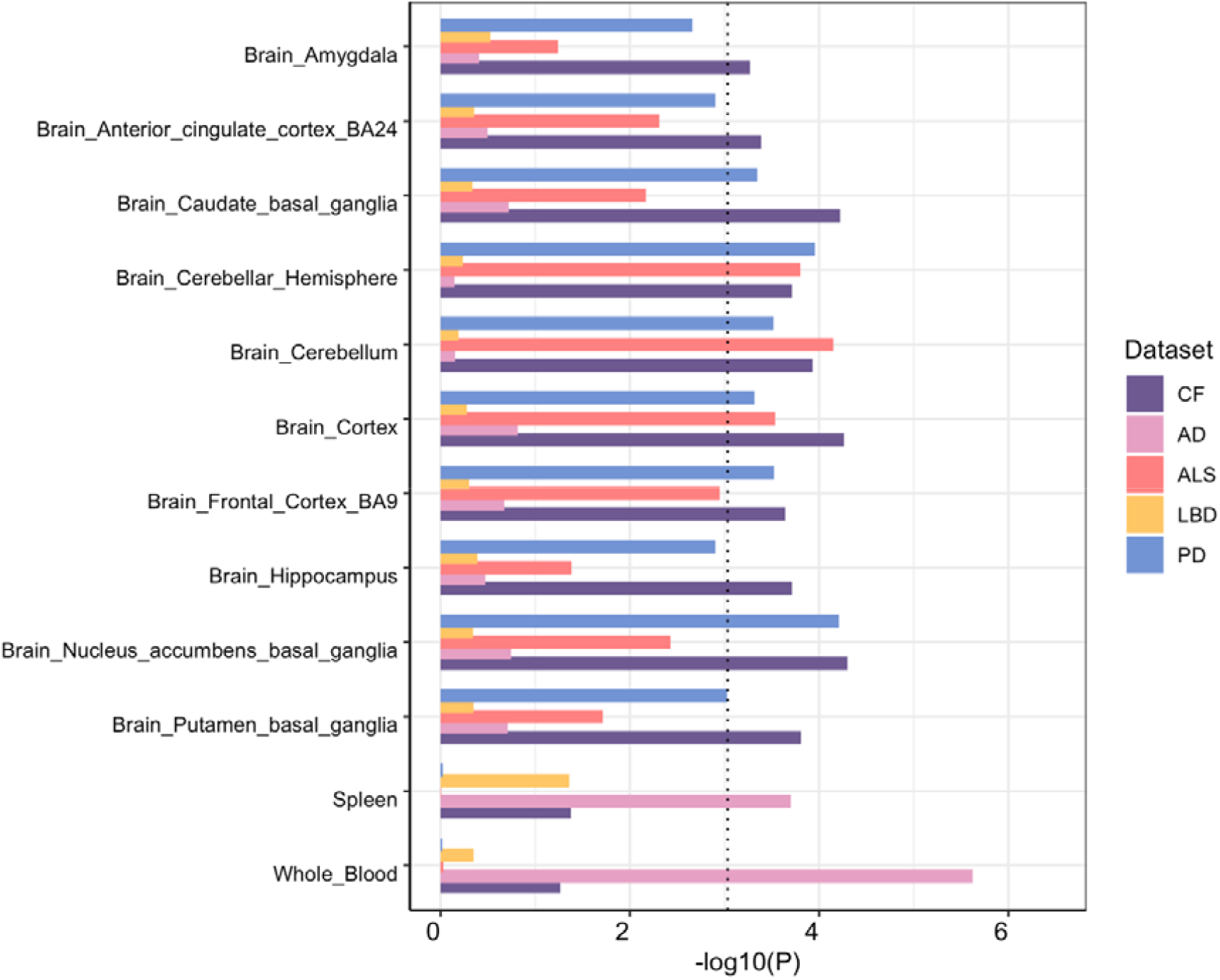
The significance values of GTEx detailed tissues that were identified as significant after Bonferroni correction for 54 tissues shows that the brain tissue significance in the common factor was largely driven by ALS and PD. The dashed line represents the Bonferroni significance threshold (0.05/54=9.26×10^−4^).

To highlight specific cell types relevant to neurodegenerative diseases, we performed FUMA celltype analysis to look for enrichment of common factor association signal in genes expressed in specific cell types. We tested all adult brain and blood tissues present in FUMA (v1.3.8) and identified microglia as the only cell type associated with the common factor after Bonferroni correction for 199 tested cell types (PsychENCODE: P=2.80×10^−5^). This cell type was significant in the AD analysis (PsychENCODE: P=2.17×10^−6^), but PD was the only other input trait where this cell type had nominal significance (PsychENCODE: P=0.0082). The results suggests that the common factor captured the AD association with microglia rather than any shared evidence for microglia. The results from all cell type analyses are available in **Supplementary Table 8**.

We also performed MAGMA gene-set analysis using the 15,496 gene-sets from MSigDB v.7.0 which were included in FUMA (v1.3.8). Eleven gene-sets were significantly associated with the common factor after Bonferroni correction for the 15,483 gene-sets which contained at least one variant (**Figure 6**; **Supplementary Table 9**). All (four) of the gene-sets significantly associated with PD and 11 of the 19 gene-sets significantly associated with AD were not significantly associated with the common factor (**Figure 6**). Three gene-sets were only significantly associated with the common factor; however, two of these were borderline significantly associated with AD. After forward conditioning, four of the eleven gene-sets significantly associated with the common factor were identified as conditionally independent, three of which have been previously associated with AD in a study^18^ which contained all of the AD data included in this study. The remaining conditionally independent gene-set (go_immune_response_regulating_cell_surface_receptor_signaling_pathway) has not been previously associated with AD and was not significantly associated with AD in this study.

**Figure 6:**
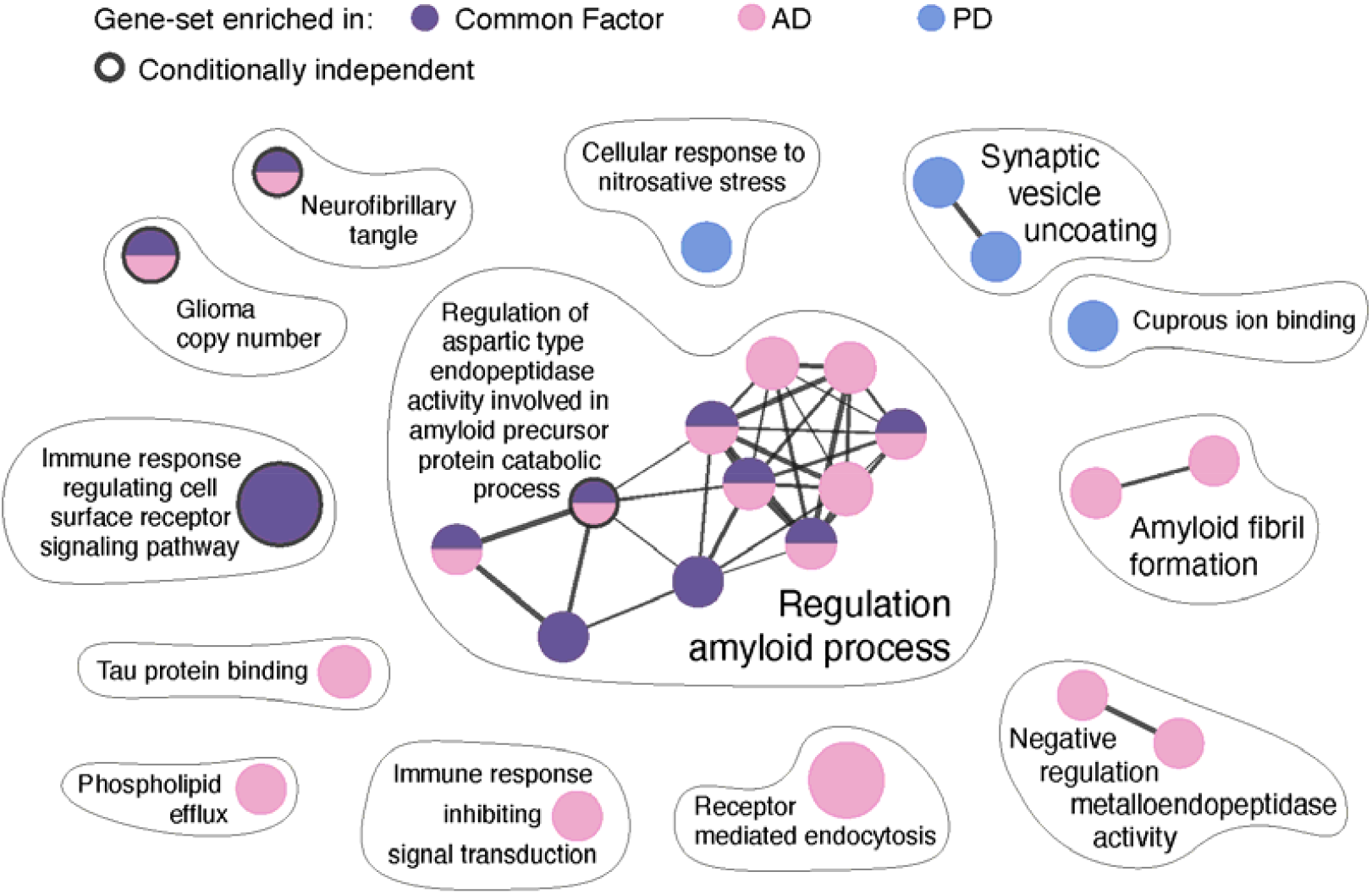
Biological process gene-set analysis results highlight the contribution of AD associated variants in the gene-sets associated with the common factor. The size of nodes represent gene-set size and edges represented similar pathways, using stringent pathway similarity scores (Jaccard and overlap combined coefficient = 0.6 as used in Paczkowska et al. (2020)^46^). The circles representing gene-sets associated with the common factor which were conditionally independent after forward conditioning are surrounded with bold outlines.

This gene-set appears to be driven by AD and LBD, with not even nominal significance in ALS and PD (P_CF_=2.91×10^−6^, P_AD_=8.42×10^−4^, P_ALS_=0.50, P_LBD_=0.0049, P_PD_=0.16). Of the 487 genes that make up this gene-set (**Supplementary Table 8**), the AD input data had 16 genes with a moderate P-value (P<1×10^−5^), where LBD only had one (*PVRL2*= 4.21×10^−10^), a gene located in the *APOE* region. The gene-sets associated with the common factor appear to be largely driven by AD genetics with only one gene-set suggesting any overlap. The one gene-set suggesting overlap had some association with AD and LBD and appears to be driven by the shared *APOE* locus.

### LAVA analyses

We used LAVA, a tool for local genetic correlation analysis, to quantify local genetic correlations between the 4 input traits and test the relationships observed from the common factor analysis. We defined local regions using the suggestive loci (P<1×10^−5^) boundaries identified from the common factor by FUMA. We also included the 4 regions identified as suggestive in two input traits but not identified as suggestive in the common factor (**Supplementary Note**). This resulted in 47 loci, with 45 loci having enough variants for local genetic correlation testing. We only performed local genetic correlation tests between traits at loci where the traits had significant local genetic heritability estimates at the locus. First, we identified loci with significant univariate heritability after Bonferroni correction for 45 loci. We identified 74 heritability estimates significantly different from 0 across 40 loci across the 4 traits (**Supplementary Figure 2**; **Supplementary Table 10**). We were then able to test 45 local genetic correlations across 24 loci where at least two traits had significant local heritability estimates. We identified 6 local genetic correlations across 4 loci that were significantly different from 0 (**Supplementary Figure 3**; **Supplementary Table 11**) after Bonferroni correction for 45 tested local genetic correlations (P<0.011).

We identified significant positive genetic correlations between ALS and PD, and LBD and PD, at the *TMEM175* locus (**Figure 3**). The local genetic correlation at this locus between ALS and LBD was also positive and nominally significant (ρ=0.62, P=0.0024). The AD data had a local heritability estimate significantly different from 0 at the *TMEM175* locus, but did not have a significant genetic correlation with any other trait. We also identified significant positive genetic correlations between AD and ALS, and AD and PD, at the HLA locus. The local genetic correlation at this locus between ALS and PD was also positive and nominally significant (ρ=0.40, P=0.0026). LBD did not have a local heritability estimate significantly different from 0 at the HLA locus. We identified a significant positive genetic correlation between AD and LBD at the *BIN1* locus, PD also had a local heritability estimate significantly different from 0 but did not have a significant genetic correlation with AD or LBD. We also identified a significant positive genetic correlation between AD and ALS at the *TNIP1* locus, PD had a local heritability estimate significantly different from 0 but did not have a significant genetic correlation with AD or ALS.

These analyses support the role of the *TMEM175* region in ALS, LBD, and PD and the role of the HLA region in AD, ALS, and PD. However, no region had significant local genetic correlations between all 4 input traits at nominal significance, which suggests no region contributes to all 4 neurodegenerative diseases. Interestingly, the *SNCA* locus was identified as significant in LBD and PD and suggestive in AD, but in the LAVA analysis no significant local genetic correlations between any of these traits were identified for the *SNCA* region. Unexpectedly, only one nominally significant correlation was identified and this was a negative correlation between AD and PD (ρ=-0.35, P=0.027). This suggests the shared association of the *SNCA* locus may be through separate mechanisms or that the linkage disequilibrium structure of this region differed between the input traits.

To further test the three-way relationships, we performed conditional analysis on the two regions with three-way relationships, conditioning on the third trait not included in the most significant local genetic correlation. The local genetic correlation between ALS and PD at the *TMEM175* locus was no longer significant after conditioning on LBD (ALS∼PD: ρ=0.77, P=2.18×10^−7^; ALS∼PD+LBD: ρ=0.38, P=0.068). No other genetic correlations at the *TMEM175* locus between two of these three traits were significant after conditioning on the third, suggesting that the genetic correlation between ALS, LBD, and PD at the *TMEM175* locus was shared (**Supplementary Table 11**). The genetic correlation between AD and ALS at the HLA locus was not significant after conditioning on PD (AD∼ALS: ρ=0.61, P=8.09×10^−5^; AD∼ALS+PD: ρ=0.26, P=0.0097). No other genetic correlations at the HLA locus between two of these three traits were significant after conditioning on the third, suggesting that the genetic correlation between AD, ALS, and PD at the HLA locus was shared (**Supplementary Table 11**). In order to prioritise a specific gene in the *TMEM175* locus, we performed local genetic correlation analysis between ALS and PD conditioned on eQTL data in that locus. However, no eQTL dataset could explain the genetic correlation between ALS and PD at the *TMEM175* locus (**Supplementary Note**).

## Discussion

With the common factor model, we aimed to identify overlapping regions, genes, gene-sets, cell types, and tissues between the 4 neurodegenerative diseases which are supported by shared genetic variants. However, we were unable to identify any region, gene, gene-set, cell or tissue type that was shared between all 4 neurodegenerative traits. This suggests that any genetic risk factors shared by AD, ALS, LBD, and PD, if present, have small effects that our study was not powered to observe, are tagged by rare variants not included in our analysis of common variants, or aggregate in gene-sets, cell types, and tissue types not tested in our analyses. We further aimed to identify novel relationships between traits at these levels that could not be identified without performing the common factor analysis. We were able to identify four brain tissues that were associated with the common factor but not any of the input traits; however, after conditional analysis we discovered that the association was with brain tissue in general, so these findings cannot be interpreted as novel. We identified three gene-sets which were only associated with the common factor; however, after conditional analysis and investigation of the genes supporting the gene-set analysis results, we suggest that these results were largely driven by AD genetics and the shared *APOE* region between AD and LBD.

We identified 4 loci which were suggestively associated with the common factor but not any of the input traits (**Supplementary Note**). These loci were investigated further using LAVA; three of these four loci did not have heritability estimates significantly different from 0 in two or more input traits, and the other locus did not show a significant local genetic correlation between the two traits with significant local heritability. This suggests that these loci are not shared. We identified one region (*DOC2A*) which had four genes significantly associated with the common factor but was not implicated in any of the 4 input traits through significant (or suggestive) genes or variants. There was some (non-significant) association for these four genes with both AD and LBD, which suggests that this region may be a general dementia region. As a whole the common factor analysis was able to identify all of the genetic overlap between the 4 input traits that could be observed through simple comparisons but was not able to go beyond this to identify additional genetic overlap.

Arneson *et al*. (2018)^5^ found no genes associated with AD, ALS, and PD; however, the HLA locus was significantly associated with these traits in the datasets used in our analyses. The identification of the HLA region as a region of interest to ALS was first reported in the GWAS catalog in 2021 (van Rheenen *et al*. (2021)^11^), four years after the analysis (September 2017) of Arneson *et al*. (2018). We were able to identify the overlap at the HLA region between these three traits because the data from van Rheenen *et al*. (2021)^11^ was included in our study. Arneson *et al*. (2018)^5^ also identified fewer overlapping genes and gene-sets between ALS and the other two traits, than between AD and PD. We replicated that finding, with the *MAPT, SNCA*, and HLA regions being shared in AD and PD, but only the *TNIP1* and HLA regions being shared in AD and ALS, and the *TMEM175* and HLA regions being shared between ALS and PD. Arneson *et al*. (2018)^5^ highlighted vesicle mediated transport as a gene-set related to AD, ALS, and PD; however, none of the gene-sets with titles containing ‘vesicle mediated transport’ had a significant association with any of the input traits or common factor in this analysis. We identified similar global genetic correlation estimates between AD and ALS and PD and ALS as reported in van Rheenen *et al*. (2021)^11^. We were unable to replicate their finding of *TSPOAP1-AS1* as a shared locus between AD and ALS but we were able to replicate their finding that the HLA and *GAK/TMEM175* loci were shared between ALS and PD. The data used in van Rheenen *et al*. (2021) overlapped extensively with the data used in this analysis. On the global genetic correlation level, the results suggest that AD and ALS are more genetically correlated than AD and PD; however, this was not reflected as a higher number of shared associated loci.

We were able to replicate the results in Guerreiro *et al*. (2016)^15^, where we also found a significant global genetic correlation between AD and DLB/LBD, DLB/LBD and PD, but not AD and PD. The genetic correlation estimates were larger in our study (AD∼DLB: rg=0.93, SE=0.28; PD∼DLB: rg=0.63, SE=0.17) compared to Guerreiro *et al*. (2016)^15^ (AD∼DLB: rg=0.58, SE=0.075; PD∼DLB: rg=0.36, SE=0.11). The differences in these estimates may be due to larger standard errors in our study and the use of different phenotypes (Guerreiro *et al*. (2016) investigated DLB, a subtype of LBD). Interestingly, there were strong global genetic correlations between AD and LBD and PD and LBD, but not a global genetic correlation significantly different from 0 between AD and PD. Despite this lack of correlation on a global level, we were able to identify some local genetic correlations between AD and PD. We replicated the finding from Stolp Andersen *et al*. (2022)^16^, where we found a significant local genetic correlation between AD and PD at the HLA region. We were also able to replicate the overlap between LBD and AD at the *APOE* and *BIN1* loci and the overlap between LBD and PD at the *GBA, TMEM175*, and *SNCA* loci identified in Chia *et al*. (2021)^14^. Overall, we were able to replicate previous positive findings and find additional overlap compared to previous studies, likely due to the increased sample size of the input datasets in our analyses. All of the previous studies mentioned in this section used datasets which were included in our analysis, so our findings cannot be considered an independent replication of previous findings.

Our study’s scope was to identify genetic overlap between all 4 input traits and was thus limited in the ability to find other relationships by the use of a common factor model. Further exploration of three- or two-way relationships would benefit from other model specifications in genomicSEM (e.g. 2 latent variables) or analysis with tools specifically designed for subset identification (ASSET^24^). Our ability to identify genetic overlap between the four traits was also limited by the relatively low heritability of the input traits (<0.1), the low genetic correlation between PD and AD and ALS, and by the relatively low sample size of the LBD dataset. As such, there is likely to be further genetic overlap between these traits than observed in this study and our study design may be useful when applied to future versions of these GWAS that have higher sample sizes. Similarly, we can only observe shared genetic overlap with the common factor when variants are present in all of the input GWAS, so some shared variants may be lost due to different GWAS study designs resulting in different sets of tested variants. Additionally, we lost 690 variants due to failed analyses of these variants; however, all of these variants were in loci that were significant or suggestive in the common factor analyses, so overlap at these loci was still examined in this study. It is possible that the genetic overlap between AD and LBD could be over-estimated due to phenotype misspecification in the original GWAS, as dementias can be difficult to differentially diagnose^25^. Using data from studies with more stringent case definitions would be beneficial to further investigate the relationship between LBD and AD, however, these studies are likely to have smaller sample size and would be less powered to identify overlapping loci. Despite these limitations, our study successfully quantified the global and local genetic overlap between four neurodegenerative diseases, AD, ALS, LBD, and PD, and highlighted two regions that overlapped between three of the diseases.

## Methods

### Data Description

We used data from 4 studies to perform the common factor analyses and the LAVA analyses. The AD data was obtained from Wightman *et al*. (2021)^18^, this data consisted of summary statistics from an inverse-variance weighted meta-analysis of all of the cohorts in their analysis except the UK Biobank and 23andMe data (39,918 cases and 358,140 controls). All individuals included in the meta-analysis were of European ancestry. The ALS data was obtained from van Rheenen *et al*. (2021)^11^ (27,205 cases and 110,881 controls) via the GWAS catalog (GCST90027164), we only used the results from the European ancestry meta-analysis. The LBD data was obtained from Chia *et al*. (2021)^14^ (2,591 cases and 4,027 controls) via the GWAS catalog (GCST90001390), the summary statistics were lifted over from GRCh38 to GRCh37 using the UCSC liftover tool^26^ to match the other data. The PD data was obtained from Nalls *et al*. (2019)^13^ (15,056 cases, 12,637 controls, 18,618 proxy cases, and 436,419 proxy controls), this data did not include the Nalls *et al*. 2014, the 23andMe post-Chang *et al*. 2017 or the Web-Based Study of Parkinson’s Disease (PDWBS) data. After downloading, all datasets were restricted to variants with MAF >0.01 and annotated with rsIDs.

### LDSC Analyses

LDSC regression^21^ (https://github.com/bulik/ldsc) was used to estimate heritability and genetic correlation estimates using HapMap3 variants only. Precalculated LD scores for LDSC were derived from the 1KG European reference population (https://data.broadinstitute.org/alkesgroup/LDSCORE/eur_w_ld_chr.tar.bz2). Population and sample prevalences were specified in the heritability and genetic correlation estimation analyses. The population prevalences for AD, LBD, and PD were the same as used in the original papers from where the data was obtained (0.05, 0.001, and 0.005 respectively). The prevalence of ALS was estimated as 0.0000625 by taking the median between the range estimated in Longinetti and Fang *et al*. (2019)^27^ (4.1-8.4 per 100[000). We used effective sample size (4*proportion of cases*(1-proportion of cases)*N) as the sample size in the munging step. Proxy cases and controls were considered the same as cases and controls when calculating the effective sample size for the PD data. The sample prevalences were set to 0.5 because effective sample size was used instead of sample size. The sample size specified (362647.1) to obtain the LDSC intercept for the common factor summary statistics was calculated by summing the effective sample sizes of the 4 input datasets. The heritability estimates of the input traits were compared to the estimates from the original studies, the PD heritability estimate was also compared to the GWAS Atlas estimate which was derived from a preprint version of the study, where the data included the proxy dataset (https://atlas.ctglab.nl/traitDB/4167).

### genomicSEM Common Factor Model

After the datasets had been restricted to common variants and annotated with rsIDs, we used genomicSEM^19^ (GitHub commit 66c1751) to munge the data. We used effective sample size as the sample size in the munging step. We then used genomicSEM to perform LD score regression on the munged data to estimate the genetic relationships between the traits. The population and sample prevalences were the same as described in the LDSC analysis. The LDSC results were then used for model estimation without variants. We specified a common factor model with LBD residual variance set to 0. The model was also specified so that the variance of the common factor was 1. We then prepped the summary statistics of the four input traits using the sumstats() function in genomicSEM. The resulting summary statistics of the four input traits were used to estimate the variant association with the common factor.

### Genomic Loci Definition

Genomic loci were defined at two levels, significant loci and suggestive loci, for all 4 input traits and the results from the common factor analysis. The significant loci were defined using default settings in FUMA (v1.3.8)^28^ whereas the suggestive loci were defined using default settings in FUMA except the maximum P-value of lead SNPs was set to 1×10^−5^. The loci were defined using LD information from the 1KG^29^ EUR data. Independent significant SNPs were defined as variants with a P-value less than the maximum P-value of lead SNPs and in low LD (r^2^<0.6). The locus boundaries were defined by these independent significant SNPs. Loci were defined in the common factor and the four input traits, any loci that overlapped or were within 100 Kb of each other across the traits and common factor were merged into a single locus and the boundaries were extended to the range of the merged locus. The loci were assigned genes to represent the locus based on previous GWAS^11,13,14,18,23,30^.

### Gene-set and Gene Analyses

Gene-set analyses were performed in FUMA (v1.3.8)^28^ using MAGMA (v1.08)^31^. All gene-set analyses were performed on the common factor analysis results and the summary statistics from the input traits. The tissue gene-set analysis tested 54 gene-sets for enrichment of association signal. The 54 gene-sets were defined by gene-expression levels from 54 GTEx tissues^32^. Significant associations were defined by a P-value lower than the Bonferroni correction threshold (0.05/54). The cell type gene-set analysis was performed in FUMA (v1.3.8)^33^ using single cell RNA sequencing data define gene-sets. We tested 199 cell types across 12 datasets, selecting only datasets from adult human brain or blood cell types (PsychENCODE_Adult^34^, Allen_Human_LGN_level1^35^, Allen_Human_LGN_level2^35^, Allen_Human_MTG_level1^35^, Allen_Human_MTG_level2^35^, DroNc_Human_Hippocampus^36^, GSE104276_Human_Prefrontal_cortex_all_ages^37^, GSE67835_Human_Cortex^38^, GSE89232_Human_Blood^39^, Linnarsson_GSE101601_Human_Temporal_cortex^40^, Linnarsson_GSE76381_Human_Midbrain^41^, PBMC_10x_68k^42^). Significant associations were defined by a P-value lower than the Bonferroni correction threshold (0.05/199).

The MSigDB v.7.0^43^ gene-set analysis tested 15,483 gene-sets for enrichment in association signal. Significant associations were defined by a P-value lower than the Bonferroni correction threshold (0.05/15483). Conditional gene-set analyses were performed by forward conditioning until no gene-sets were significant. Initially, the significant gene-sets were conditioned on the most significant gene-set, then the gene-sets were conditioned on the most significant gene-set from the initial analysis and the most significant gene-set from the first round of conditioning. This process was repeated, with further gene-sets added for each round of conditioning, until no gene-set was significant. The final set of gene-sets which were used for conditioning were the conditionally independent gene-sets. MAGMA gene analyses were also performed using FUMA (v1.3.8) with default settings. Significant associations were defined by a P-value lower than the Bonferroni correction threshold for the number of genes tested (CF=18248; AD=19065; ALS=18554; LBD=18657; PD=18912). All significant gene-sets were visualized as a graph in Cytoscape^44^ using EnrichmentMap^45^. The size of nodes represented gene-set size and edges represented similar pathways, using stringent pathway similarity scores (Jaccard and overlap combined coefficient = 0.6 as used in Paczkowska *et al*. (2020)^46^).

### LAVA Analyses

The LAVA^20^ analyses were performed to identify local genetic correlations between specific regions. The AD, ALS, LBD, and PD summary statistics, after restriction to common variants (MAF>0.01) and annotation with rsID, were used as input for the LAVA analyses. The LAVA (v0.0.7; GitHub commit 0dd05b6) analyses used a sample overlap estimate (genetic covariance intercept) from the LDSC analyses to adjust for potential sample overlap (**Supplementary Table 12**). As with the genomicSEM analyses, effective sample size was used instead of sample size. Local heritability estimates were calculated for all 4 of the input traits across the merged suggestive loci (loci suggestive in the common factor or suggestive in two or more input traits). The merged suggestive loci boundaries were defined previously by FUMA (v1.3.8) (see **Methods: Genomic Loci Definition**). Local observed heritability estimates were converted to the liability scale using formula 23 from Lee *et al*. (2011)^47^ and the population prevalances from the genomicSEM analysis. Any regions with two or more traits with local heritability significantly different from 0 after Bonferroni correction for 45 loci (0.05/45) were tested for genetic correlation between the traits with significant heritability at that locus. Significant genetic correlations were defined by a P-value lower than the Bonferroni correction threshold for the number of genetic correlations tested (0.05/45).

Conditional analyses were performed using the run.pcor() function in LAVA (v0.07). For the conditional analyses, the pairwise significant genetic correlations at the loci with 3 significantly correlated traits were conditioned on the remaining significantly genetically correlated trait. This was performed for all 3 possible conformations. To test whether the genetic correlation between ALS and PD at the *TMEM175* locus could be explained by eQTL data, we performed genetic correlation analysis between the two traits at this locus conditioned on eQTL data. In order to find suitable eQTL data, we downloaded 19 brain or neuron datasets from the eQTL catalogue^48^. We included the following datasets: Braineac2_ge_putamen^49^, Braineac2_ge_substantia_nigra^49^, BrainSeq_ge_brain^50^, CommonMind_ge_DLPFC_naive^51^, GTEx_ge_brain_amygdala^32^, GTEx_ge_brain_anterior_cingulate_cortex^32^, GTEx_ge_brain_caudate^32^, GTEx_ge_brain_cerebellar_hemisphere^32^, GTEx_ge_brain_cerebellum^32^, GTEx_ge_brain_cortex^32^, GTEx_ge_brain_frontal_cortex^32^, GTEx_ge_brain_hippocampus^32^, GTEx_ge_brain_hypothalamus^32^, GTEx_ge_brain_nucleus_accumbens^32^, GTEx_ge_brain_putamen^32^, GTEx_ge_brain_spinal_cord^32^, GTEx_ge_brain_substantia_nigra^32^, ROSMAP_ge_brain_naive^52^, Schwartzentruber_2018_ge_sensory_neuron^53^. We trimmed the data to the *TMEM175* region (GRCh38: chr4:822968-1036991) then lifted over the data to GRCh37, and annotated it with rsIDs. We split the data so that each dataset contained eQTLs for a single gene and then identified eQTL-gene pairs with significant local heritability after Bonferroni correction for 978 eQTL-gene pairs (0.05/978). We then estimated the local genetic correlation between the significant eQTL-gene pairs with ALS, LBD, and PD, assuming no sample overlap between the eQTL data and the input traits. After Bonferroni correction for 246 genetic correlation tests, there was a single eQTL-gene pair that was significantly correlated with 2 of the traits (ALS and PD). We then conditioned the local genetic correlation between ALS and PD on the significant eQTL-gene pair to test whether the eQTL-gene pair dataset could explain the association between ALS and PD at the *TMEM175* locus.

## Data Availability

The summary statistics from the common factor model analysis and AD summary statistics will be made available at https://ctg.cncr.nl/software/summary_statistics after publication. The summary statistics for the ALS, LBD, and PD can be obtained from their original publications: Nalls et al. (2019) https://doi.org/10.1016/s1474-4422(19)30320-5 Chia et al. (2019) https://doi.org/10.1038/s41588-021-00785-3 van Rheenen et al. (2021) https://doi.org/10.1038/s41588-021-00973-1.

## Data Availability

The summary statistics from the common factor model analysis and AD summary statistics will be made available at https://ctg.cncr.nl/software/summary_statistics after publication. The summary statistics for the ALS^11^, LBD^14^, and PD^13^ can be obtained from their original publications.

## Code Availability

The code used in these analyses will be made available at https://github.com/dwightman/Neurodegeneration after publication.

## Acknowledgements

DP was funded by The Netherlands Organization for Scientific Research (NWO VICI 453-14-005), NWO Gravitation: BRAINSCAPES: A Roadmap from Neurogenetics to Neurobiology (Grant No. 024.004.012), and a European Research Council advanced grant (Grant No, ERC-2018-AdG GWAS2FUNC 834057). DW was funded by NWO Gravitation: BRAINSCAPES: A Roadmap from Neurogenetics to Neurobiology (Grant No. 024.004.012). IEJ was funded by NWO Gravitation: BRAINSCAPES: A Roadmap from Neurogenetics to Neurobiology (Grant No. 024.004.012). JES was supported by funding from the Amsterdam Neuroscience Alliance Project. CR was funded by NWO Gravitation: BRAINSCAPES: A Roadmap from Neurogenetics to Neurobiology (Grant No. 024.004.012). Analyses were carried out on the Genetic Cluster Computer hosted by the Dutch National computing and Networking Services SurfSARA.

## Author contributions

DPW analysed the data. DPW wrote the manuscript. DPW, IEJ, JES, CR, ET, and DP designed the analysis plan. IEJ, JES, and DP supervised the project.

