## Supplementary Note for "The genetic overlap between Alzheimer’s disease, amyotrophic lateral sclerosis, Lewy body dementia, and Parkinson’s disease"

Suggestive loci

To investigate whether there was further sub-threshold overlap between AD, ALS, LBD, and PD, we repeated the FUMA significant loci definition (**Results; Methods**), except we allowed for loci to be defined around variants with suggestive P-values (P<1x10^-5^). There was a total of 269 suggestive loci across the common factor and the 4 input traits (**Supplementary Table 3**), 43 were suggestive in the common factor, 101 were suggestive in AD, 67 were suggestive in ALS, 32 were suggestive in LBD, and 80 were suggestive in PD. In addition to the relationships identified through loci defined with significant variants (P<5x10^-8^), *TMEM175* was suggestive in ALS and significant in LBD, PD, and the common factor, *SNCA* was suggestive in AD and significant in LBD, PD and the common factor, and *MAPT* was suggestive in AD and significant in PD and the common factor. At this level, the HLA locus, the *SNCA* locus, and *TMEM175* locus were all at least suggestive in three of the four traits; however, no locus was suggestive in all four traits.

In contrast to the analysis at the significant level, not all loci suggestively associated with the common factor were suggestive in at least one of the input traits, and not all loci suggestive in two or more input traits were suggestive in the common factor. Four loci were only suggestive in the common factor, and four loci were suggestive in two or more input traits but not suggestive in the common factor. Of the four common-factor-only loci, two (*SHARPIN* and *TPCN1*) are loci which overlap with loci identified in a recent AD GWAS^18^. Visual inspection of the four loci that are suggestive in two input traits but not in the common factor suggests that these loci may be separate loci in relative proximity to each other.

We looked at the univariate heritability and genetic correlation of the loci identified as only suggestive in the common factor and the loci which were suggestive in two or more input traits but not the common factor (**Supplementary Table 13**) to determine whether these regions were truly shared regions. Of the four loci only associated with the common factor, only one locus (*PLEC*) had two traits with local heritability significantly different from 0 (AD: h2_liability_=0.0013, P=4.04x10^-7^; PD: h2_liability_=6.18x10^-4^, P=6.18x10^-5^). However, the genetic correlation between AD and PD at this locus was not significantly different from 0 (ρ=0.44, P=0.64). One locus (*EFL1/CPEB1*) that was suggestive in two input traits but not the common factor also had significant univariate heritability in two input traits (AD: h2_liability_=0.0011, P=8.97x10^-5^; ALS: h2_liability_=3.88x10^-4^, P=5.77x10^-6^). However, this locus was not even a nominally significant correlation between the two traits (ρ=0.262, P=0.24). This suggests that the suggestive loci only identified in the common factor and the suggestive loci identified in two or more input traits but not the common factor are unlikely to be regions of true overlap.

TMEM175 locus eQTL conditioning analysis

In order to prioritise a specific gene in the *TMEM175* locus, we performed local genetic correlation analysis between ALS and PD conditioned on eQTL data of that locus. Initially, we ran LAVA univariate h2 analyses for all eQTL-gene pairs in the *TMEM175* locus from the 19 brain and neuron eQTL datasets available in the eQTL catalogue^20^. This identified 82 eQTL-gene pairs with a local observed heritability significantly different from 0 after Bonferroni correction for 978 eQTL-gene pairs. Then we tested for local genetic correlations between these 82 eQTL-gene pairs with ALS, LBD, and PD resulting in 3 significant local genetic correlations after Bonferroni correction for 246 local genetic correlations. Of these 3 significant local genetic correlations (**Supplementary Table 11**), two were the same eQTL-gene pair (*SLC26A1* in GTEx_ge_brain_hippocampus) correlated with ALS and PD. We then conditioned the genetic correlation between ALS and PD on the *SLC26A1*-GTEx_ge_brain_hippocampus data, which only slightly decreased the significance of the genetic correlation between ALS and PD (ALS~PD: ρ=0.77, P=2.18x10^-7^; ALS~PD+eQTL: ρ=0.85, P=1.28x10^-5^). This result suggests that we were unable to identify an eQTL and gene pair that could explain the association between ALS and PD at this locus.

Supplementary Figures

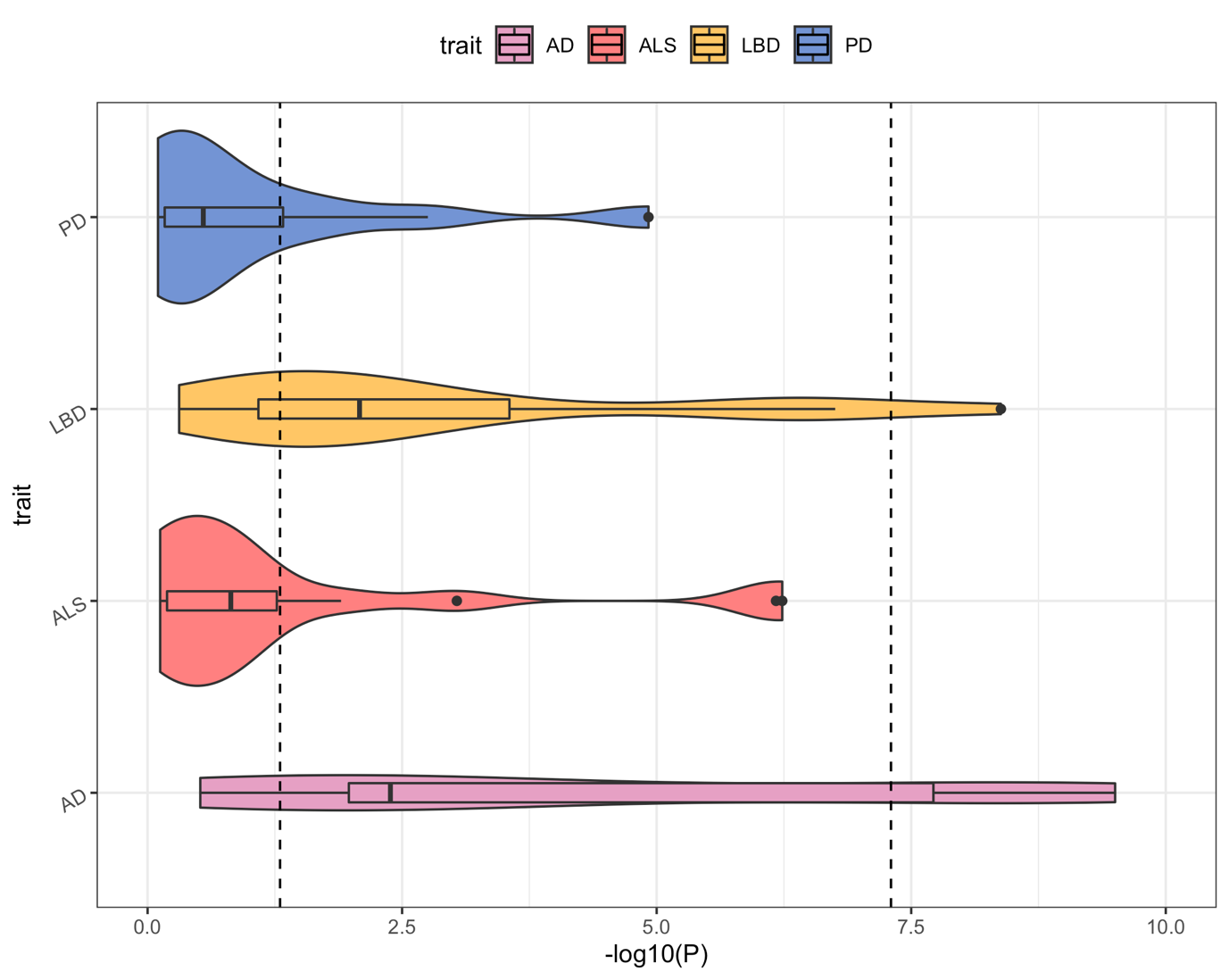

Supplementary Figure 1: The P-values of the common factor lead variants in the 4 input traits identifies AD and LBD as having smaller P-values in the common factor lead variants. The first dashed line represents the nominal significance threshold (0.05) and the second dashed line represents the Bonferroni corrected significance threshold (5x10^-8^). Box plots are within the violin plots, the box displays the first quartile, the median value, and the third quartile. The lines out from the box plot end at the first quartile minus 1.5 times the interquartile range and the third quartile plus 1.5 times the interquartile range. Dots represent P-values of variants where the P-values are outside of the first quartile minus 1.5 times the interquartile range and the third quartile plus 1.5 times the interquartile range.

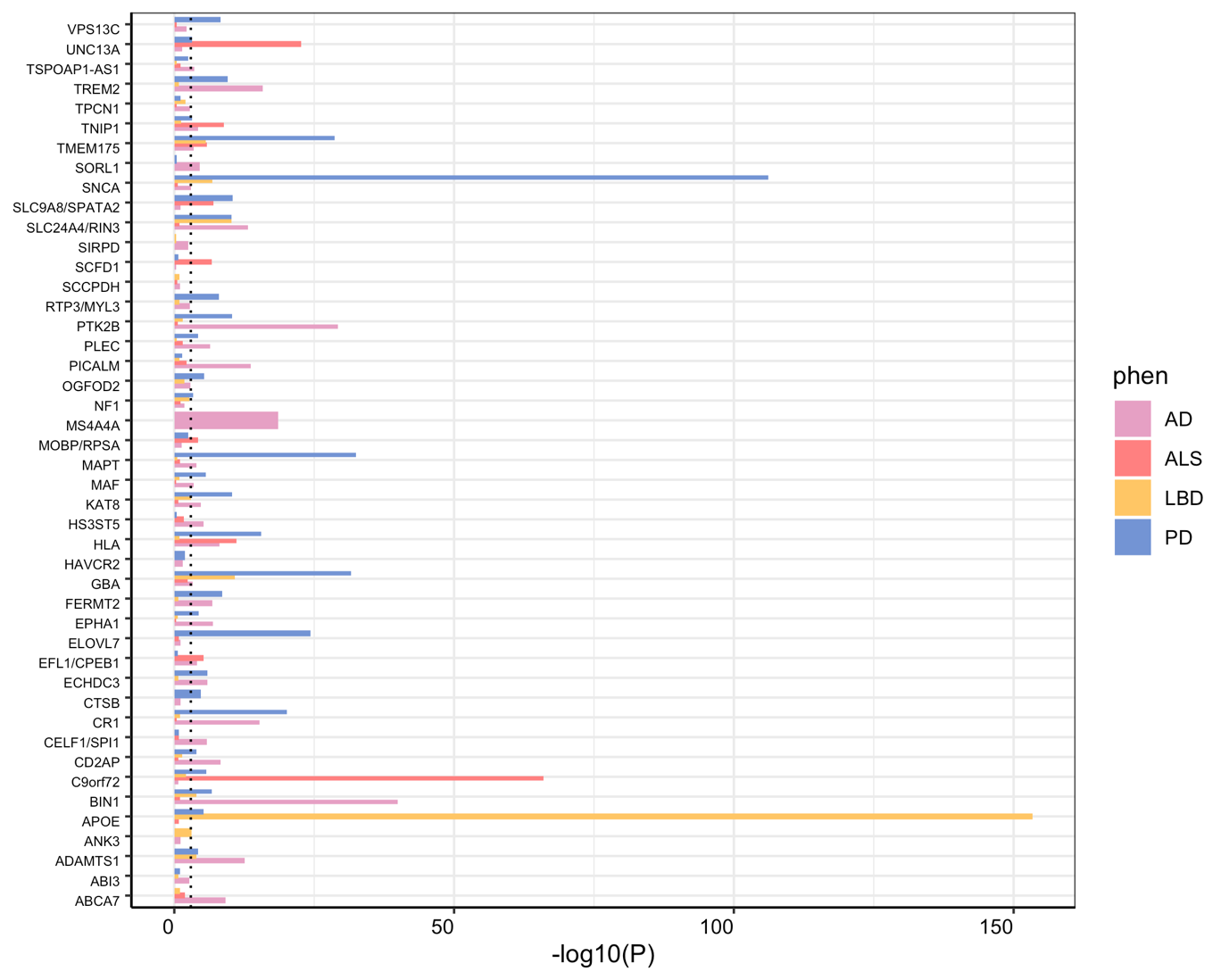

Supplementary Figure 2: The P-values of the local heritability estimates for each of the 4 input traits across the 45 loci show traits where the local heritability is significantly different from 0. The dashed line represents the Bonferroni corrected significance threshold (0.05/45 loci). Heritability estimates that could not be estimated due to lack or excess of association signal are not included in the plot.

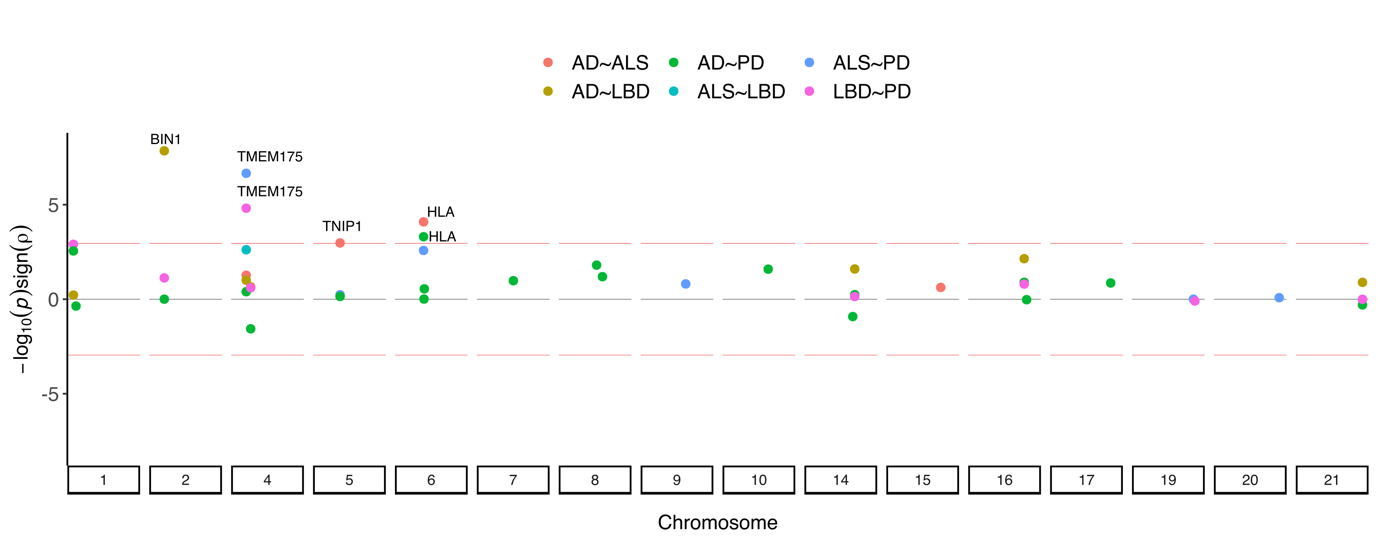

Supplementary Figure 3: The bivariate genetic correlation estimates between traits at loci where these traits had significant local heritability highlights 4 regions with significant genetic correlations after Bonferroni correction for 45 genetic correlations tests. The red lines represent the Bonferroni correction threshold (0.05/45 genetic correlations) multiplied by the direction (sign) of the genetic correlation.
